# Development of the Individual Participant Data (IPD) Integrity Tool for assessing the integrity of randomised trials using individual participant data

**DOI:** 10.1101/2023.12.11.23299797

**Authors:** KE Hunter, M Aberoumand, S Libesman, JX Sotiropoulos, J Williams, W Li, J Aagerup, BW Mol, R Wang, A Barba, N Shrestha, AC Webster, AL Seidler

## Abstract

**Introduction:** Increasing concerns about integrity in medical research have prompted the development of tools to detect untrustworthy studies. Existing tools focus on evaluating aggregate or published data, though some trustworthiness issues may only be detected upon scrutiny of individual participant data (IPD). To address this, we developed the IPD Integrity Tool for detecting integrity issues in randomised controlled trials with IPD available. This manuscript describes the development of this tool.

**Methods:** We conducted a literature review to collate and map existing integrity items. These were discussed with an expert advisory group, and agreed items were included in a standardised tool and automated where possible. We piloted this tool in two IPD meta-analyses, and conducted preliminary validation checks on 13 datasets with and without known integrity issues in a blinded manner.

**Results:** The literature review identified 120 integrity items: 54 could be conducted at the publication or aggregate data (AD) level, 48 required IPD, and 18 were possible with aggregate data, but more comprehensive with IPD. Based on these items, an initial reduced tool was developed in a consensus process involving 13 advisors with different backgrounds (countries, profession, education). This initial tool included 11 items across four domains for AD, and 12 items across 8 domains requiring IPD. The tool was iteratively refined throughout piloting on two IPD meta-analyses including a total of 116 trials (73 with IPD, and 43 with only AD available), and preliminary validation using an additional 13 datasets. All five studies with known integrity issues were accurately identified during validation. The final version of the tool included seven domains with 13 items for AD and eight domains with 18 items requiring IPD.

**Conclusions:** The quality of evidence informing health care relies on trustworthy data. This manuscript describes the development of a tool to enable researchers, editors, and other stakeholders to detect integrity issues in randomised trials using IPD. Detailed instructions on the application of this tool will be published subsequently.

## BACKGROUND

There are increasing concerns within the research community that some published studies are based on low-quality, falsified, or fraudulent data. The traditional peer review system focuses on identifying the novelty, utility, and methodological robustness underpinning research findings. This process assumes that all researchers are acting in good faith and is inadequate to confirm the veracity of data. Failure to filter out untrustworthy studies that are plagued by errors or bias (whether intentional or not) can distort the evidence base which underpins guidelines and clinical practice, contributing to potential research waste, patient harm and general mistrust in research. Therefore, it is crucial to ensure that data are genuine and to assess the integrity of studies^1^ before relying on their results.

### What is research integrity?

Research integrity can be defined as adherence to the highest professional standards and ethical principles in conducting research^2,3^ (‘best practice’, Figure 1). This instils trust and confidence in the methods and findings of a study^4^ and centres around embedding practices that enhance the quality, reliability and relevance of research.^5^ At the other end of the spectrum is worst practice or deliberate misconduct, which comprises fabrication (making up data or results), falsification (manipulating research materials, images or data to misrepresent research), and plagiarism. Collectively, these are often referred to as FFP (fabrication, falsification, plagiarism). Questionable research practices (also termed irresponsible or detrimental research practices) comprise a broad category that fills the gap between FFP and best practice; they may be intentional or inadvertent (honest errors, ignorance). Although considered less serious than FFP, questionable research practices are widely prevalent and are suggested to be a much larger contributor to research waste and potential patient harm than FFP.^2^ Questionable research practices are many and varied, but may include authorship breaches, poor study design, poor study conduct, data dredging, inappropriate statistical analyses, selective reporting and improper handling and reporting of missing data.^6,7^

**Figure 1.**
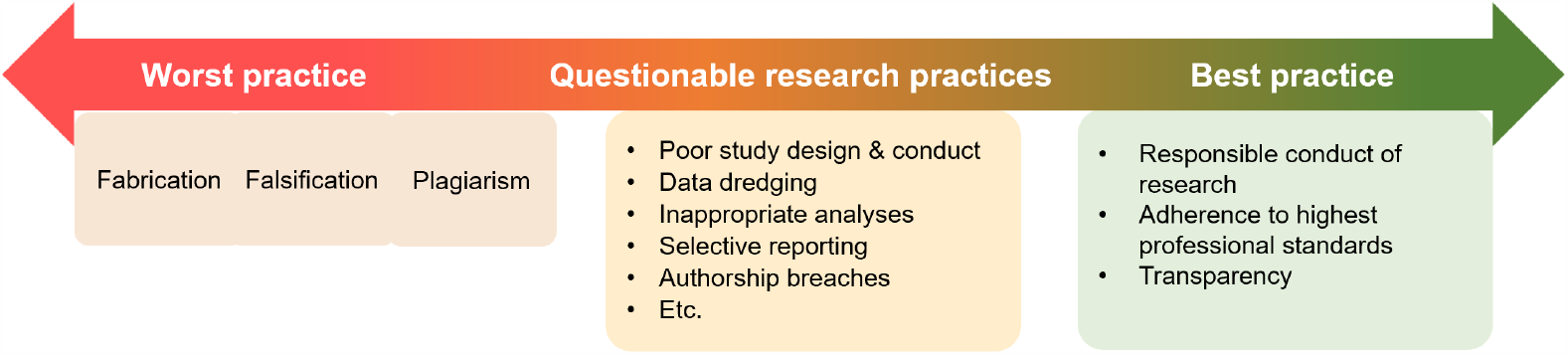
Research integrity continuum *(adapted from Steneck)*^*2*^

### What has been done to date?

Concerns about the integrity and trustworthiness of medical research have prompted calls for resources and guidance to support editors, publishers, researchers and other key stakeholders to detect and manage potentially fraudulent, misleading or low-quality studies.^8-10^ In response, there have been an increasing number of publications offering guidance and suggesting methods to assess integrity of studies. For instance, Weibel et al^9^ developed a research integrity assessment (RIA) tool for assessment of randomised controlled trials of investigational medicinal products (IMPs) for evidence synthesis. The primary purpose of the tool is to identify potentially problematic studies (i.e. studies not in accordance with good clinical practice) by evaluating six domains, including retraction notices, prospective trial registration, ethics approval, authorship, reporting and plausibility of methods, and plausibility of results. Bordewijk et al.^11^ also published a comprehensive scoping review in which they identified 27 methods to assess research misconduct in health research, categorised into overall, textual, image and data concerns. More recently, Parker et al.^8^ conducted a qualitative study drawing on the experiences of experts in the field to identify warning signs of research fraud and misconduct. Cochrane are developing a tool to identify problematic RCTs in systematic reviews of health interventions (INSPECT-SR),^12^ and have released a collaboration-wide policy for managing potentially problematic studies in systematic reviews,^13,14^ while the Cochrane Pregnancy and Childbirth Group have developed and begun implementing their own trustworthiness screening tool.^15^ REAPPRAISED^16^ is also a useful list to guide scrutiny of published articles, and the Trustworthiness in RAndomised Clinical Trials (TRACT) checklist has recently been published^10^ to help reviewers triage RCTs based on the risk of fabricated data. Domains covered by TRACT include governance, author group, plausibility of intervention usage, timeframe, drop-out rates, baseline characteristics, and outcomes. The issue of plagiarism has already received widespread attention, and there are now effective tools for its detection.^11,17^ However, these may need to be re-visited as the development and use of artificial intelligence systems such as ChatGPT are rapidly evolving.^18^

### Remaining gaps

While these resources are important contributions to the field, there are some limitations and remaining gaps. First, existing tools focus on assessing published studies or summary-level/aggregate data, rather than individual participant data (IPD), i.e. raw line-by-line data for each participant. Yet, IPD are often required to detect trustworthiness issues. This is evidenced by about a 20-fold increase in the detection of false trial data in studies submitted to *Anaesthesia*, from 2% when IPD were not available up to 44% when they were.^19^ Consequently, experts have argued that we can no longer trust aggregate data, and that evidence synthesists should request and personally review the veracity of IPD for all studies before including them in meta-analyses.^20^ Second, there is a lack of practical advice on how to apply suggested integrity checks and tools, and what constitutes an untrustworthy study.^8^ This introduces an element of subjectivity and uncertainty about what action to take if any issues are identified. Third, a common criticism of integrity checks is that they are seen to be overly burdensome or requiring specific expertise, suggesting a need for education and automation.^8,9,11^

## AIM

We aimed to develop a user-friendly individual participant data (IPD) integrity tool for assessment of the integrity and trustworthiness of randomised controlled trials.

## METHODS

We adapted approaches for the development of checklists and measurement tools recommended by Moher et al^21^ and Streiner et al^22^ to develop the tool. This was a four-step process including: i) defining the scope and making preliminary conceptual decisions; ii) literature review and item generation; iii) expert advisory group consultation; and iv) piloting, validation, and refinement of the tool.

### i) Defining the scope and making preliminary conceptual decisions

This stage involved consideration of the primary purpose of the integrity tool, in what circumstances it may be applied, and any important characteristics that are required or preferred.

### ii) Literature review and item generation

We sought to identify existing integrity and trustworthiness items to assess studies and their raw data. First, we extracted information from a recent scoping review^11^ and references cited in this review. Additionally, we conducted searches of Google Scholar, identified data integrity items from our prior experience conducting several IPD-MA and by attending relevant conferences and webinars.

### iii) Expert advisory group consultation

Throughout development of the tool, we consulted an international expert advisory group, comprising 13 members from Australia, the United Kingdom and Europe. This multidisciplinary group had diverse expertise as methodologists, clinicians, systematic reviewers, data managers, consumers, and statisticians.

### iv) Piloting, preliminary validation and refinement of tool

We pilot tested the tool in two IPD meta-analysis projects, one on cord management in preterm infants,^23-25^ and the other on initial oxygen in preterm newborns.^26^ Each eligible trial was assessed by two independent reviewers using the tool and any potential issues identified were elaborated on in a comments field.

To assess preliminary sensitivity and specificity, the tool was tested by a blinded reviewer in a sample of 13 RCTs with IPD datasets available: five that were known to be problematic (confidentially sourced from journal editors), and eight without known issues sourced from online publications or data repositories. Datasets were ordered using a random sequence generator, and the reviewer was asked to check only the IPD domains first, since the aggregate data domains could compromise blinding, e.g. if a publication was retracted. The reviewer was only given information on the participants, interventions, comparators, and outcomes to enable assessment of the dataset. No identifying data or information that could compromise objectivity or introduce bias were provided (e.g. author name, country, journal). Once IPD checks were complete, the reviewer conducted aggregate data checks. Although the reviewer remained blinded to whether this was a previously identified untrustworthy study, they were provided citations for any publications at this stage, since this information is required for these checks.

Throughout piloting and validation, the tool was iteratively refined based on advisor feedback.

## RESULTS

The results of each step in developing the IPD Integrity Tool are displayed in Figure 2 and elaborated below.

**Figure 2.**
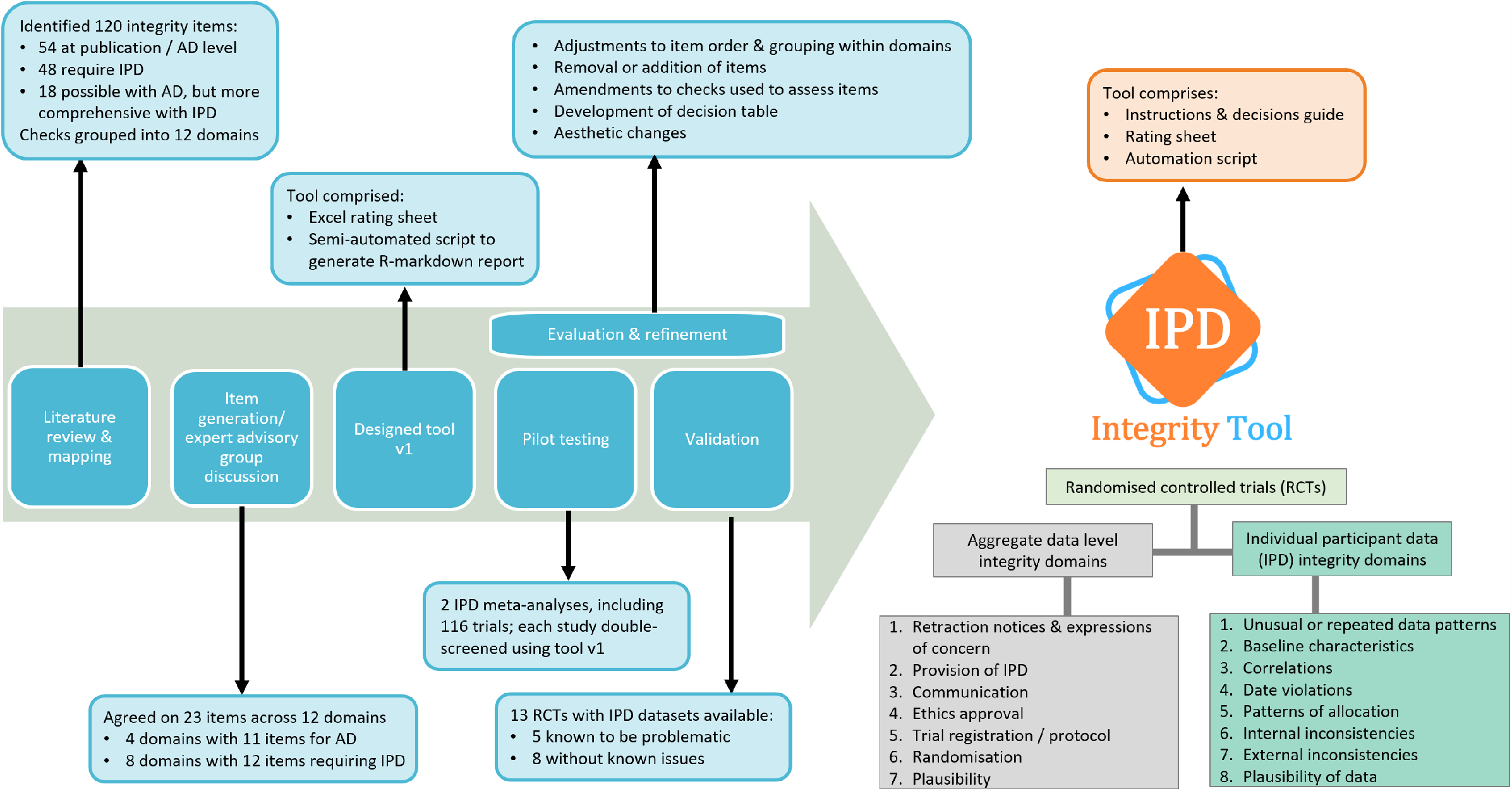
Development of the IPD Integrity Tool

### i) Defining the scope and making preliminary conceptual decisions

This tool was initially designed for the assessment of the integrity and trustworthiness of randomised controlled trials for inclusion in IPD meta-analyses. Key specifications were that the tool should: i) identify untrustworthy studies and provide guidance on how to manage these; ii) be user-friendly; iii) enable high inter-rater reliability; and iv) enable clear reporting of results of integrity assessments. Based on advisor feedback, external interest and need, we decided to broaden the scope of the tool to editorial processes, so that it may be applied to important scenarios beyond IPD meta-analysis, as part of routine editorial checking processes or to aid journal editors with investigation of questionable studies.

### ii) Literature review and item generation

The literature review identified 120 existing integrity items (Appendix 1). These were grouped into 12 key domains and categorised as either assessable at publication or aggregate data level (n=54), requiring IPD (n=48), or possible with aggregate data but more comprehensive with IPD (n=18).

### iii) Expert advisory group consultation

Identified items were discussed among an international multidisciplinary expert advisory group for potential application to a large IPD meta-analysis. After review and discussion, the advisory group agreed on a total of 23 items across 12 domains. This included four domains with 11 items for aggregate data and eight domains with 12 items requiring IPD. Agreed items were incorporated into a standardised tool, which included detailed instructions on how each item should be assessed, how to rate each item (no issues, some/minor issues, or many/major issues), interpretation of results, and practical guidance on the appropriate course of action. Where possible, checks were automated to improve efficiency and make the process less resource intensive. This was done using the R Markdown in R version 4.1.3,^27^ which enabled data, R code, plots, tables, and written notes to be combined into a single reproducible report.^28-30^

### iv) Piloting, preliminary validation and refinement of tool

#### Piloting

This stage involved applying the tool to two IPD meta-analysis projects in neonatology, including a total of 116 trials. Of these, 73 trials provided IPD, and for the remaining 43 only aggregate data were available from publications. The tool identified at least one potential integrity issue in 57 of the 73 (78%) trials providing IPD. Most were minor or genuine errors that were resolved via consultation with trial investigators, resulting in more complete and high-quality datasets contributing to the meta-analysis. Three of the 73 trials providing IPD were excluded from meta-analyses due to integrity concerns, including major discrepancies between IPD and published data, and lack of association between variables known to be highly correlated.

As expected, trials that did not provide IPD raised more serious trustworthiness concerns. Of the 43 studies for which only published aggregate data were available, nine (21%) were excluded due to multiple concerns regarding trial registration, randomisation, ethics, lack of communication and plausibility.

Where applicable, the collective pattern of potential integrity issues identified for a trial was discussed among our advisory group. This enabled us to develop and pilot decision rules to guide exclusion of trials with integrity issues.

During piloting, we considered whether publication in predatory journals should be an item on our integrity tool. However, most evidence synthesis experts surveyed on this topic^31^ agreed that the most important consideration is the quality and validity of the study and its results, rather than where it is published, and therefore studies published in predatory journals should not be automatically excluded from evidence syntheses

#### Preliminary validation

Of the 13 datasets, the reviewer accurately excluded all five of the studies with known issues after completing the IPD checks, giving 100% sensitivity. Of the eight datasets without known integrity issues, all were judged sufficiently trustworthy upon completion of all checks.

#### Evaluation and refinement

Based on feedback and evaluation throughout piloting and preliminary validation, and in consultation with the advisory group, the tool was iteratively refined to make it more intuitive, logical, and user-friendly. Most refinements were minor adjustments to item order and collapsing/expanding groupings of items or domains. Substantive changes included removal of one item that was deemed less useful (‘Inspection of inliers’), and addition of a new item (‘Plausibility of author group’). Some items remained the same, but the method of assessment was changed or enhanced, e.g. ‘Excessively homogeneous distribution of binary baseline variables’ was initially tested by comparing expected and observed numbers of consecutive pairs of events in the dataset, but changed to a simple *Runs test*,^32^ and Pearson correlation coefficients were added to correlation graphs for ease of interpretation. Several amendments were also made to the R-markdown script, including the addition of an instructive vignette, and tidying of the output.

### The final tool

The final tool includes seven domains with 13 items for aggregate data and eight domains with 18 items requiring IPD (Figure 2). The Tool comprises an instructions and decision guide which explains how to assess and rate each item, a rating sheet to indicate whether there are no issues, some/minor issues, or many/major issues for each item, and a template R script to semi-automate assessment of some items. Detailed guidance on application of the Tool is available elsewhere.^33^

## DISCUSSION

We developed the IPD Integrity Tool to screen for potential integrity issues in randomised controlled trials using individual participant data. The Tool includes seven aggregate data domains and eight domains specific to IPD. The development process involved a literature review, consultation with an expert advisory group, piloting, validation, evaluation, and refinement.

The main goal of this tool is to ensure that data contributing to the evidence base are reliable and trustworthy, so that patients may receive the best care. This tool should not be used for ‘witch-hunting’ or to raise anxiety among researchers who act in good faith. We note that some items in the tool overlap or interplay with risk of bias and data cleaning procedures, and IPD-specific methods to streamline these processes are in development.^34^

### Strengths

To our knowledge, this is the first integrity tool that focuses on examining raw line-by-line IPD. This enables increased detection of trustworthiness issues, by allowing more comprehensive quality and integrity checks to be undertaken than those based on aggregate data alone, e.g. scrutiny of data patterns and correlations. The tool was developed based on comprehensive literature searches, and in close consultation with an international advisory group comprising methodologists, clinicians, systematic reviewers, data managers, consumers, and statisticians. It was pilot tested and validated in a range of datasets, and iteratively refined and improved based on evaluation.

### Limitations

Validation of integrity tools such as this one is challenging since there is no agreed gold standard for the assessment of performance accuracy. We used datasets that had been investigated and found to have known integrity issues for validation, together with datasets that had no known integrity issues. Yet, it is possible that the latter were untrustworthy trials. Further, categorisation of a study as trustworthy or not may be subject to a spectrum of opinion.^19^ Our preliminary validation dataset was small. In future, we aim to further validate this tool.

A commonly raised concern is that by publishing tools like this one, we may assist fabricators to adapt their techniques to avoid detection.^8,10^ Conversely, improved detection of integrity issues may deter undesirable research behaviours.^35^ Regardless, techniques for checking integrity need to evolve to keep pace with the latest questionable research practices and misconduct, as well as methods to circumvent our data checks. We intend to update the tool to maintain currency by scoping emerging practices.

### How can we better protect research integrity?

More proactive approaches are required among the research community to prevent integrity problems from arising.^36^ This requires a shift in focus from the ‘publish or perish’ mindset, where publication pressure is associated with a higher prevalence of questionable research practices,^37^ to prioritising and institutionalising responsible research methodology and practice.^7,38^ Further, journal editors and publishers have a responsibility to uphold integrity by ensuring that they only publish research that has been conducted in accordance with internationally accepted guidelines.^39^ To achieve this, they should routinely screen manuscripts for integrity issues,^40^ and we support arguments to require sharing of raw data prior to publication.^10,31^ Among journal readers and reviewers, a healthy scepticism is also encouraged; while publications were commonly accepted as truth in the past, as Moore et al argue “the biggest mistake of all, then, is taking evidence on trust and without checking it.”^41^

In future, we aim to explore how this tool might be adapted and applied to non-randomised and observational studies, as well as emerging novel designs like umbrella and basket trials. In addition, rapid advancements in artificial intelligence and statistical simulation pose an increasing threat to data integrity and must be explored further.

## CONCLUSION

It is important to check the veracity of a study’s IPD before relying on its results. We have developed the IPD Integrity Tool to enable researchers, editors, publishers, and other interested stakeholders to screen randomised controlled trials for integrity issues.

## Data Availability

All data produced in the present study are available upon reasonable request to the authors.

## Acknowledgements

The authors are grateful to our international expert advisors, including Lisa Askie, Lelia Duley, Alan Montgomery, Jack Wilkinson, and Gill Gyte.

