## Appendix 1 for "Development of the Individual Participant Data (IPD) Integrity Tool for assessing the integrity of randomised trials using individual participant data"

**Appendix 1. Integrity / trustworthiness checks identified in literature review**

? indicates check could be done at some level using AD, but deeper examination is possible with IPD

| **Item** | **Aggregate data/**  **Publication/study-level** | **Individual participant data (IPD)** |
| --- | --- | --- |
| **Domain: Randomisation** | | |
| 1. Non-random sequence generation patterns - assess using statistical tests (e.g. runs test^1^ or tests within the R package ‘randtests’)^2,3^ |  | 🗸 |
| 2. Randomisation inadequacy (allocation concealment)^2,3^ | 🗸 |  |
| 3. Plot cumulative accrual for each group over time^4,5^ |  | 🗸 |
| 4. Non-uniform distribution of day of randomisation across groups^6^ |  | 🗸 |
| 5. Unexpected days of randomisation (Sundays, holidays)^6,7^ |  | 🗸 |
| 6. Large differences in number of participants per group (compared to pre-specified allocation ratio) | 🗸 |  |
| 7. Unexpectedly even/exactly equal number of participants across groups (within trials) and block randomisation was not used^2,8,9^ | 🗸 |  |
| 8. Unexpectedly even number of participants randomised across trials by same authors and block randomisation was not used^2^ | 🗸 |  |
| **Domain: Baseline characteristics** | | |
| 9. No or few baseline characteristics reported (particularly key prognostic characteristics) | 🗸 |  |
| 10. No or few baseline characteristics provided in IPD dataset (particularly key prognostic characteristics) |  | 🗸 |
| 11. Baseline characteristics excessively/improbably similar between randomised groups within trials, e.g. groups have (near) identical values for age, ethnicity, sex^2,8-17^ | ? | 🗸 |
| 12. Baseline characteristics excessively/improbably similar between randomised groups across trials by same authors^2,9^ | 🗸 |  |
| 13. Baseline characteristics excessively different between randomised groups, e.g. major imbalances between groups in age or sex^9-11,18,19^ | ? | 🗸 |
| 14. Baseline imbalance in the outcome variable^20,21^ | ? | 🗸 |
| 15. Baseline p value distribution deviates from expected distribution^12-17,19,22-26^ for group means (continuous data, using t-tests/ANOVA) or proportions (categorical data, using Chi-square test or Fisher’s Exact) for all randomised participants. Can use Monte Carlo simulations / Carlisle’s method^3,27^ (but note some issues)^16,23^ and/or Kolmogorov-Smirnov test^28,29^ | ? | 🗸 |
| 16. Excessively homogeneous distribution of binary baseline variables^19^ |  | 🗸 |
| 17. Significantly different variance of continuous baseline variables between groups^19^ |  | 🗸 |
| **Domain: Data patterns / variable distributions** | | |
| 18. Digit preference (first or leading digit) / Benford’s law^7,26,30-36^ (Note: only use when first digit can range between 1 & 9). Benfords and digit pref program’ available at <http://www.ctc.ucl.ac.uk/Training.aspx> | ? | 🗸 |
| 19. Digit preference (last or terminal digit):^7,9,16,18,19,26,34-41^ Compare last digit frequency distribution to a known distribution (generally uniform) OR Compare mean and SD of last digit value within a participant to study-wide distributions using CI approach   R-program available in van den Bor^36^ <https://github.com/chartgerink/ddfab> | ? | 🗸 |
| 20. Round number preference/excess of round numbers^7^  - R-program: integer_check^35^ - ‘integers program’ available at <http://www.ctc.ucl.ac.uk/Training.aspx> | ? | 🗸 |
| 21. Zipf’s law^42^ |  | 🗸 |
| 22. Outliers (too few or too many) using graphical methods (e.g. boxplot), descriptive statistics, Cook's distance, Mahalanobis Distance (large MD = outlier^41^), Grubb’s method, Euclidean distance, and/or R program outlier_check^2,6,7,10,20,21,35,39^ |  | 🗸 |
| 23. Multivariate inliers^7,37,39^     i. Mahalanobis distance (MD): compare between each participant vs study-wide value for key variables. Small MD = inlier^21^ |  | 🗸 |
| 24. Internally inconsistent, impossible, or illogical data,^10^ e.g. age cannot be negative, a person who has died cannot have a heart rate |  | 🗸 |
| 25. Sequences, duplication or groupings of data values, patterns, repetition within columns and rows that are extremely unlikely to have occurred by chance^17,19,43^     i. Carlisle^17^ methods: Format IPD spreadsheet in Excel, colour cells by value & duplication to highlight outlying values, patterns & repetition within columns & rows. Plot column values overall & by group. Calculate probability of sequence similarity among groups through simulation, resampling values & calculating the sum of differences between values in the same position of the sequence in each group. Use resampling to calculate probability of runs of the same value in columns. Analyse sequences of modified values, e.g. after deleting the integer to the left of decimal place & analyse sequences of values right of the decimal place |  | 🗸 |
| 26. Expected correlations between variables are not present, are too weak or too strong, or are in the wrong direction.^7,21,26,35-37,41,44^ Can be assessed at participant level, site level and study level (where applicable).     i. Calculate Pearson correlation (e.g. using ‘Proc Freq’ in SAS)^41^     ii. Angular clustering^44^     iii. Neighbourhood clustering^44^     iv. ‘Correlation check’ available at <http://www.ctc.ucl.ac.uk/Training.aspx>     v. R-program available in van den Bor^36^ | ? | 🗸 |
| 27. Multivariate associations that are extremely different to those estimated from control data that are (arguably) genuine^26^ |  | 🗸 |
| 28. Repeated measures:     i. Possible interpolation, duplicates or invented patterns, e.g. assess using autocorrelations, profiles, polynomial contrasts, runs tests^7,45^     ii. carryover effect = exact match of a value for a participant from one visit to next, even though change is expected^39,41^     iii. repeated values = number of identical values for participant within visit/overall^41^     iv. Repeated measurements clustering (Euclidean distance, Mahalanobis distance)^44^     v. Variances in biological variables surprisingly consistent over time^7,10,21,45^ |  | 🗸 |
| 29. Abnormally low within person variability^20,21,45^     i. Check SD distributions across visits for each participant; compare study-wide to participant-specific confidence intervals^41^ |  | 🗸 |
| 30. Data errors^2,10,45,46^ | ? | 🗸 |
| 31. Large deviations between observed and expected distributions for continuous and categorical variables^47^     i. expected sampling variability is known through Central Limit Theorem.^24^     ii. Construct histograms of standardised mean difference^21,47^     iii. Significantly greater clustering of standardised mean differences around zero than would be expected by chance (for continuous variables, the SDs should be ∼1.0 )^47^ |  | 🗸 |
| 32. Strange peaks in variable distribution^7^ |  | 🗸 |
| 33. Data too skewed (single variables)^7^ |  | 🗸 |
| 34. Granularity-related inconsistency of means (GRIM) test (for discrete data)^48^ – looks for mathematically or clinically impossible values of means and SDs^49^ | 🗸 |  |
| 35. Variability of reported measures mathematically impossible (GRIMMER test)^50^ | 🗸 |  |
| 36. Plausibility of standard deviations, e.g. too similar to be plausible^21,26,51,52^ | ? | 🗸 |
| 37. Plausibility of coefficients of variation, e.g. too similar to be plausible^51,53^ | ? | 🗸 |
| 38. Too little or too much variance^7,24,35,45^ - compare observed vs expected distribution of variances^26^     i. R-program: variance_check^35^     ii. ‘Variance check’ available at <http://www.ctc.ucl.ac.uk/Training.aspx> |  | 🗸 |
| 39. Values too close or too far from the means^20,35^ |  | 🗸 |
| 40. Hotelling T^2^ to check several variables at a time^7^ |  | 🗸 |
| **Domain: Results** | | |
| 41. Impossible or conflicting results / internal inconsistency,^10^ e.g. more ongoing pregnancies than clinical pregnancies | ? | 🗸 |
| 42. Implausible results – consider magnitude of estimated effect, positive outcomes, consistency with literature and/or biology^2,8,9,21,26,29,54,55^ | ? | 🗸 |
| 43. Highly unusual outcome frequency without reasonable explanation (particularly for rare outcome)^10^ | ? | 🗸 |
| 44. Underreporting of adverse events, especially in high risk participants (e.g. multiple comorbidities) and across centres for multicentre trials^41^ | 🗸 |  |
| 45. Summary outcome data excessively similar or identical across study groups^10,19^ | 🗸 |  |
| 46. identical or highly similar outcome values across trials by same author^16,28,29^ | 🗸 |  |
| 47. Reduced variability of results^20,21^ | 🗸 |  |
| **Domain: Missing data** | | |
| 48. High proportion of missing data^10^ | ? | 🗸 |
| 49. Missing or duplicate participant IDs (if sequential)^5,56^ |  | 🗸 |
| 50. Check if data appear to be included for all randomised participants,^5^ (as per any publications, registration records, Consort diagram, etc.) |  | 🗸 |
| 51. Missing data appear too perfect across groups or are implausibly few.^51^     i. Calculate rate of missing data by dividing the number of missed measures by the number of times it was expected.^41^ | ? | 🗸 |
| 52. (Close to) zero or unrealistically low losses to follow up (consider whether compatible with disease, age, timeline^2,8-10,29^ | 🗸 |  |
| 53. Losses to follow-up result in perfectly rounded number in each group (e.g. 50 or 100) | 🗸 |  |
| 54. Participant withdrawals not reported^3^ | 🗸 |  |
| **Domain: Dates** | | |
| 55. Impossible, illogical or infeasible order of dates^7,19,35,36^     i. R-program: date_order_check^35^     ii. Residual plots^7^     iii. CUSUM Control charts,^7^     iv. date order program’ available at <http://www.ctc.ucl.ac.uk/Training.aspx>) |  | 🗸 |
| 56. Routine measurements taken at weekends or public holidays^20,35,37,41^  - weekends (R-program: weekend_hol_check (option: weekends)^35^ - public holidays (R-program: weekend_hol_check (option: holiday)^35^ - ‘Program instructions - 2 weekends and national holidays’ available at [www.ctc.ucl.ac.uk/Training.aspx](http://www.ctc.ucl.ac.uk/Training.aspx) - ‘R-program available in van den Bor^36^ |  | 🗸 |
| 57. Check distribution of visit lag, i.e. distance of actual visit date relative to the target date, on site and participant levels (where applicable)^41^ |  | 🗸 |
| 58. Check if ordering and spacing of tests meets study guidelines^37^ |  | 🗸 |
| 59. Reported follow-up duration is inconsistent with recruitment dates and manuscript submission date | 🗸 |  |
| **Domain: Recruitment** | | |
| 60. Implausible / infeasible recruitment timeframe (consider disease epidemiology, sample size, stringency of eligibility criteria, study location)^2,7-10,29,36,54,55^ - R-program available in van de Bor^36^ | 🗸 |  |
| **Domain: Consistency cross-checks between IPD, associated publications and registration records** | | |
| 61. Discrepancies between IPD (or reported data) and eligibility criteria specified in publication(s) and/or registration record(s ^10^ | ? | 🗸 |
| 62. Inconsistencies in the distribution of baseline characteristics between publication and IPD^57^ |  | 🗸 |
| 63. Inconsistencies in enrolment dates between IPD, registration record and/or publication |  | 🗸 |
| 64. Inconsistencies in overall participant numbers and participant numbers per group, e.g. between CONSORT diagram, registration record, publication text/tables, and IPD^57^ |  | 🗸 |
| 65. Other external inconsistencies between publication, registration, IPD, e.g. sample size, location, results, study methods^2,10^ |  | 🗸 |
| 66. IPD not provided for relevant variables/outcomes that have been reported in publication and/or registration record |  | 🗸 |
| 67. Inconsistency between summary statistics (e.g. median (IQR), mean (SD)) calculated from IPD and publication^17,57^ |  | 🗸 |
| 68. Primary or other outcomes changed from registration to publication^10^ | 🗸 |  |
| **Domain: Multicentre trials / central statistical monitoring** | | |
| 69. Tests for slippage^7^ |  | 🗸 |
| 70. Chernoff faces to compare mean responses across sites^37^ or across several variables at a time^7^ |  | 🗸 |
| 71. Star (needle, spike) plots to compare mean responses across sites^37^ or across several variables at a time^7^ |  | 🗸 |
| 72. Noticeable differences in means and variances of baseline characteristics in one centre versus another^18^ | ? | 🗸 |
| 73. Extreme difference in correlation coefficients between a pair of variables within each site and/or among sites^44^ |  | 🗸 |
| 74. Large absolute difference between correlations in a centre compared to correlations for all sites combined^36^ |  | 🗸 |
| 75. Unexpected correlation of repeated measurements at each centre - calculate intraclass correlation coefficient (ICC)^38^ |  | 🗸 |
| 76. Unexpected variability of measurements among centres using folded F-test^38^ |  | 🗸 |
| 77. Unexpected distribution of leading digits from centre compared to all other sites together (Use chi-square test)^35^ |  | 🗸 |
| 78. Unexpected distribution of last digit frequency over all participants at each centre to that from all other centres:  - Fisher exact test^41^ - chi-square test^38,41^ - dissimilarity index^36^ - visual examination of plots^37^ |  | 🗸 |
| 79. Unexpected differences in day of week of randomisation across centres using Pearson chi-squared test^38^ |  | 🗸 |
| 80. Imbalanced baseline characteristics across sites  - For binary data, compare the proportion with a characteristic at each centre to the overall proportion from all other centres using a Pearson chi-squared test^38^ - For continuous variables likely to be roughly normally distributed, use t-test, using pooled standard deviation to compare mean at each centre to overall mean of other centres^38^ - Noticeable differences in means and variances in one trial versus another^18^ |  | 🗸 |
| 81. Excess of very dissimilar means^27^ at each centre compared to overall mean of other centres:^51^ Calculate a distance measure to indicate how far away one centre’s data are from the overall mean across all centres, standardised by the overall standard deviation^38^ |  | 🗸 |
| 82. Extreme differences in outcome rates across centres (adjusted for country)  - calculate the probability of observing an adjusted outcome rate as extreme as that observed at that centre, assuming a Poisson distribution with the overall adjusted mean^38^ |  | 🗸 |
| 83. Extreme differences in the proportion of missing data between centres^36^  - R-program available in van den Bor^36^ |  | 🗸 |
| 84. Principal component analysis of p-values across centres^45^ |  | 🗸 |
| **Domain: Study governance** | | |
| 85. Absent or retrospective trial registration^3,8-10^ | 🗸 |  |
| 86. Protocol not available^8,9^ | 🗸 |  |
| 87. Ethics approval issues^3,8-10^ | 🗸 |  |
| 88. Involvement of study authors in ethical oversight^2^ | 🗸 |  |
| 89. Funding source / COI not reported or implausible given resources required^2,10^ | 🗸 |  |
| 90. Study location(s) implausible based on study design and methods^10,58^ | 🗸 |  |
| 91. Refusal to share IPD^3,8,9^ | 🗸 |  |
| 92. Inadequate contact/communication with authors^9^ | 🗸 |  |
| **Domain: Publication checks – authors** | | |
| 93. Authorship feasibility/plausibility, e.g. <3 authors for large RCT^58^ | 🗸 |  |
| 94. Authors do not meet criteria for authorship^10^ | 🗸 |  |
| 95. Absence of contributorship statement^10^ | 🗸 |  |
| 96. Productivity of author team – implausible volume of work by group/authors^2,10^ | 🗸 |  |
| 97. Author history of retraction, fraud or misconduct^2,16,34,47,52,53,58,59^ | 🗸 |  |
| 98. Author profile/track record: no staff page on institution website, no other publications despite being professor – check affiliation and publication record | 🗸 |  |
| **Domain: Other publication checks** | | |
| 99. Retraction notice for study^58^ | 🗸 |  |
| 100. Published expression of concern for study^58^ | 🗸 |  |
| 101. Published in predatory journal | 🗸 |  |
| 102. Misleading text / misinformation in publication^2^ | 🗸 |  |
| 103. Implausible interval between study completion & manuscript submission^10,60^ | 🗸 |  |
| 104. Compatibility of summary data with range of possible values,^10^ e.g. SPRITE (Sample Parameter Reconstruction via Iterative Techniques)^51,61^ | 🗸 |  |
| 105. Inconsistencies within publication, e.g. methods are inconsistent with what is presented in results, discrepancies between data reported in figures, tables, text^2,3^ | 🗸 |  |
| 106. Plagiarism^10,62,63^  - HelioBLAST by HelioText plagiarism detection software (formerly known as eTBLAST)^64-66^ (<https://helioblast.heliotext.com>) - Déjà vu, duplicate citation database^65^ - Statistically Improbable Phrases (SIPs) method^67^ - iThenticate commercial plagiarism detection software and manual verification^68,69^ - Turnitin (links from scoping review): <https://www.crossref.org/services/similarity-check/>, <http://www.ithenticate.com/>, <https://www.turnitin.com/> | 🗸 |  |
| 107. Image manipulation or duplication^10,51,70-72^ | 🗸 |  |
| 108. Inappropriate statistical methods, tests and/or interpretation^2,3,10,21^ | 🗸 |  |
| 109. Compatibility of statistical test results with reported data/ Reproducibility of statistical analysis^3,10,51^  - Automated check using R program Statcheck^49,51,73,74^ <https://cran.r-project.org/web/packages/statcheck/index.html> - Recalculate test statistics: <https://cran.r-project.org/web/packages/rpsychi/index.html> | 🗸 |  |
| 110. Discrepancies between the values for percentage and absolute change^10^ | 🗸 |  |
| 111. Incorrect calculations of proportions and percentages^10^ | 🗸 |  |
| 112. Inappropriate algorithms used to derive variables or score outcomes^2^ | 🗸 |  |
| 113. Subgroup means incompatible with those for the whole cohort^10^ | 🗸 |  |
| 114. Numbers that do not add up across a table^20^ | 🗸 |  |
| 115. Numbers in graphs differ to those quoted in text^2,3,20^ | 🗸 |  |
| 116. Incorrect units reported^10^ | 🗸 |  |
| 117. Inconsistent or incorrect numbers of participants throughout publication^10^ | 🗸 |  |
| 118. P values quoted without the data necessary to estimate them^20^ | 🗸 |  |
| 119. Typographical errors^10^ | 🗸 |  |
| 120. Duplicate reporting or publication of data^2,10^ | 🗸 |  |
